# Natural Language Processing Techniques to Identify Zoonosis Awareness

**DOI:** 10.1101/2023.05.06.23289607

**Authors:** Roger Geertz Gonzalez

## Abstract

In this study, we incorporated several NLP techniques to identify the most important factors in the open-ended responses part of the *Knowledge, Attitudes, and Practices: Survey of Zoonoses in Wildlife Trade (KAP)* in Cambodia. These included: TF-IDF, ngrams, Latent Semantic allocation (LSA), k-means, Latent Dirichlet Allocation (LDA), and Top2Vec. The top topics participants identified included 1) stating that they handled wildlife by setting traps and mist nets, 2) stating they were bitten by bat or rat, 3) which zoonotic symptoms caused sickness, 4) describing how they would go to the hospital when they came down with zoonotic symptoms, and 5) saying that they were aware of avian flu and its symptoms.

Based on our findings, recommendations for Cambodian public health officials include: 1) they need to educate participants to wear protective gear to prevent from being bitten by bats and rats during their jobs with these animals, and 2) they need to educate participants about the danger of different types of zoonotic diseases including Ebolavirus, Mojianvirus, etc., so that these participants can recognize the risks when handling bats and rats, and so they can take early action by seeking medical help as soon as they are bitten.

---

TF-IDF, ngrams, LSA, k-means, LDA, and Top2Vec techniques of KAP survey in Cambodia reveal the role of bats and rats in possible zoonosis transmissions

## Introduction

Natural Language Processing (NLP) are a set of computer techniques that searches for the understanding of unstructured data (Ganegedara, 2018). Unstructured data consists of text, images, and videos (Sarkar, 2019). NLP has been used recently for disease surveillance and mitigation including: web searches for disease terms, applying text mining analysis to understand the genetic structure of SARS-CoV-2, extract top topics related to heart disease, chronic kidney disease, and dengue.

According to (Baclic et al., 2020), NLP has allowed for the rapid identification of population diseases, the identification of interventions and outcomes related to infectious disease for disease surveillance and has supported disease prevention and health promotion. Other NLP public health uses include: analysis of unstructured electronic health data, analysis of risk behaviors using social media, automated and systematic review of public health publications, and identification of promising public health interventions via peer-reviewed publications (Baclic et al., 2020). Our NLP analyses focuses on finding the underlying knowledge, behaviors and attitudes related to wildlife and zoonoses in Cambodia and to articulate best preventive practices.

We proffer specific examples of how specific NLP algorithms have been used for disease surveillance and prevention and then explain how we used each of these to analyze the *Knowledge, Attitudes, and Practices: Survey of Zoonoses in Wildlife Trade (KAP)* in Cambodia open-ended survey. These were interviews of participants in Cambodia 2016, 2018, and 2019. 1,555 KAP surveys were administered by interviewers in 98 communes, 73 of 170 districts located in 21 of the country’s 24 provinces as well as Phnom Penh (Forestry Administration, 2017). The open-ended questions focused on wildlife disease knowledge asking which diseases are transmitted by wild animals, what are some clinical signs of diseases transmitted by wild animals, and how we can protect ourselves from these diseases.

Jang et al. (021) demonstrate how Term frequency-inverse document frequency (TF-IDF) could be used to detail frequent infectious disease terms from a variety of text data on the internet. They built an automated web-based disease outbreak monitoring system using a deep-learning-based data filtering and ranking algorithm for disease-related topic extraction. One of their filters uses TF-IDF to rank disease-related hot words which included from highest to lowest ranking: “SARS-CoV 2,” “president,” “world,” “cases,” “China,” “people,” “states,” “Trump,” “infections,” and “conference.”

el Boujnouni et al. (2021) implemented ngrams to extract the sequence genomic data as texts from SARS-CoV-2 to show that its genomic data was similar to pangolins in different locations around the world. Ngrams are consecutive sequences of words that are correlated together and thus, applicable for genomic sequencing (Silge & Robinson, 2017). el Boujnouni et al. (2021) combined ngrams for text categorization, principal component analysis for dimensionality reduction, and Random Forest algorithm for supervised classification to find the origin of the SARS-CoV 2 virus by comparing its nucleic acid sequence with other coronaviruses.

Gefen et al. (2018) conducted latent semantic analysis (LSA) to extract topics related to congestive heart failure from medical records. The researchers found that “cardiac” was closely related to “body parts” and “procedures” than to “diagnoses.” They also found that “hypertension” was related to “hyperlipidemia” (high levels of fat particles in the blood) and in general, to co-occurring conditions and diagnoses. LSA is a valuable tool because it shows related terms that medical professionals might miss during a cursory reading.

K-means has been used in several health settings including research on Alzheimer’s and dementia (Alashwal et al., 2019). Luong & Chandola (2017) used k-means to group patients suffering from chronic kidney disease based on their disease progression profiles. Violán et al. (2018) used k-means in conjunction with multiple correspondence analysis to identify multimorbidity patterns from patients’ electronic health records.

Topic modeling has been used to understand the relationship between infectious diseases. In their review of SARS-CoV-2, MERS, and SARS research publications, Cheng et al. (2020) found that “patient,” “case,” and “infection” were the top topics found in research publications about these diseases. These findings could help the medical community by highlighting useful themes and interrelationships to prevent a quick snapshot among research literature.

In their study of the initial outbreak of SARS-CoV-2 in China, Liu et al. (2020) found via Latent Dirichlet allocation (LDA) that media reports failed to capture the outbreak on time and thus, failed to become a leading indicator of the disease outbreak. While they found that the major media themes during the initial outbreak consisted of prevention and control procedures, medical treatment, and research, these were found to be overgeneralized and not refer specifically to SARS CoV-2.

Ye et al., (2016) used LDA to study social media topics about dengue in Guangdong Province in China. They found that social media posts about dengue correlated with dengue cases in the region. The top topics they found included: “prevention,” “detection,” “fear,” “symptoms,” and “care.” These were then correlated with a spatial analysis of actual cases. They suggest that social media via LDA and spatial modeling can be used to detect real-time dengue infections.

Crane et al. (2020) used LDA to identify the most current SARS-CoV 2 topics and distill Covid-19 research into specific areas. These areas could then be searched by others to find their specific areas of interests. The top ten topics they found included: “treatments,” “risk factors,” “infection,” “symptoms,” “research studies,” “transmission,” “public health caregiver risk,” and “genetics virology.”

Pietsch & Lessmann (2018) studied whether human coding of open-ended responses was better than topic modeling. They found that topic modeling has some advantages over human coding because it: 1) saves time and money, and 2) can reduce human biases. Additionally, they found that LDA is comparable in performance to other methods such as Latent Feature Latent Dirichlet Allocation, Bi-term Topic Model, and Word Network Topic Model regarding analysis of open-ended responses.

We applied NLP (TF-IDF, ngrams, LSA, k-means, LDA, and Top2Vec) techniques to the *Knowledge, Attitudes, and Practices: Survey of Wildlife Trade* in the KAP open-ended survey to determine the backgrounds of the participants regarding handling wildlife trade. We also used NLP techniques to detail their specific actions when encountering infectious disease symptoms or accidents handling wildlife. By identifying the top topics and activities related to zoonotic disease, we can make suggestions to health experts about how they could improve zoonotic disease prevention.

## Materials and Methods

The KAP survey was administered between April and October 2015 and April and November 2016 across 21 provinces of Cambodia (International Development Research Centre). In the KAP open-ended survey responses, participants were asked the following: what is your wildlife title?, where do you work?, describe an accident with wildlife you have had, what kind of protective equipment do you wear when handling wildlife?, what specific actions do you take when there is a disease outbreak?, which wildlife infectious disease are you familiar with?, which wildlife infectious disease symptoms are you aware of?, and any additional responses they would like to communicate. These 2038 participant open-ended responses in the KAP open-ended survey were transformed to a corpus which was then analyzed using NLP. All NLP analyses was done using R version 4.1.0 (R Core Team, 2020) except for the Top2Vec algorithm which was done in Python 3.6.

TF-IDF, invented separately by Hans Peter Luhn (Luhn, 1957) and Karen Spärck Jones (Jones, 1972), calculates how frequently it appears in a document (term frequency [tf]) and multiplies it with the inverse document frequency (idf) which decreases the weight for commonly used words and increases the weight for words that are used less frequently in the documents (Silge & Robinson, 2017). The TF formula is: *tf (w _1_D) = f_wd_* where f_wd_ is the frequency for word **w** in document **D**. The IDF formula is:

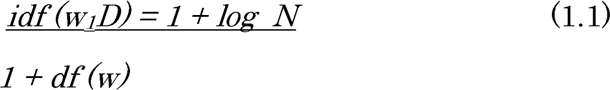

where each term is divided by the total number of documents by the document frequency for each term and then applying logarithmic scaling to the result (Sarkar, 2019). 1 is then added to the document frequency for each term to indicate that there is one more document that has every term in the vocabulary (Sarkar, 2019). We used the *tm* package (Feinerer et al., 2020) to convert the survey to a *corpus* or an object containing raw strings and for the TF-IDF, ngrams, LSA, K-Means and LDA analyses. The *tm* package allows for the important of text data, corpus handling, preprocessing and creation of term-document matrices (Feinerer et al., 2020). Preprocessing of the corpus included: a. normalizing the frequencies, stemming (converting all words to their stem form, for example, “feces” to “fece”), removing stop words or common English words such as “is,” “the,” and “want,” cutting out any words with 2 letters or less, e. removing numbers, and f. removing punctuation (Sarkar, 2019). Then the *document-term-matrix*was created which is a sparse matrix (matrix with mostly zero values) describing the corpus of documents with one row for each document and one column for each term (Silge & Robinson, 2017).

The ngram learning process includes three sub modules: a Hidden Markov Model (HMM), a sentence planner, and a discourse planner (Ganegedara, 2018). HMM is used to predict the next word given a specific sequence. It states that the probability of future states depends only on the present and not on the sequence of events that came before it (Goodfellow et al, 2016). The sentence planner corrects for any linguistic mistakes. The discourse planner correctly orders and structures a set of messages. The text corpus was converted to bigrams and trigrams by transforming the corpus document to tokens.

LSA assumes that in each document, a latent structure exists between the words that are related and combines related words together in the same space (Sarkar, 2019). LSA uses Singular Value Decomposition (SVD), a mathematical technique, used to reduce the number of rows while preserving the similarity structure between the columns in a term-document matrix. Thus, groups of related words can be summed up into distinct topics or concepts (Shmueli et al., 2020; Ganegedara, 2018). Values close to 1 are closely related terms while values close to 0 are dissimilar.

K-means is an unsupervised learning method where the data is not labeled. It focuses on clustering by dividing data into different groups. In this case, the goal is to identify meaningful groups (Bruce et al., 2020). The same document-term-matrix used above for the TF-IDF and LSA analyses was used for the k-means model. The criteria that k-means uses to identify meaningful groups is to minimize *inertia*, or within-cluster *sum-of-squares* (Sarkar, 2019). To run k-means, an initial number of centroids needs to be chose first. Two centroids were chosen initially to run the k-means model. Afterwards, an elbow plot was chosen to determine the best number of centroids to use. The elbow “joint” or cutoff for best number of centroids according to the plot was three. The k-means was then rerun with three centroids.

Text mining, like latent Dirichlet allocation (LDA) is an unsupervised learning method used to extract information or relationships from unstructured data (Sarkar, 2019). LDA uses a Bayesian probabilistic model of text documents by transforming these to a “bag of words,” a feature extraction method that discards the order or structure of words (Sarkar, 2019). It then generates topics from documents by using a probability distribution that follows a Dirichlet polynomial prior distribution. LDA is efficient because it can handle text-specific dimensions without making assumptions and it can be used with little programming (Sarkar, 2019). All the words in the document were stemmed where only the root of the words were kept to reduce noise in the data (Sarkar, 2019). Then stop words (e.g., the, to, of) (Silge & Robinson, 2017), small words, punctuation, numbers, very frequent and infrequent terms were removed. Number of topics were set to 5 to represent the number of expected topics based on the specific questions about wildlife and infectious diseases.

Another topic modeling method that was created recently in 2020 is Top2Vec. (Angelov, 2020) created Top2Vec because LSA and LDA, according to him, have several weaknesses: the number of topics have to be known, stop-words have to be listed, and stemming and lemmatization are required. With Top2Vec, these are unnecessary, and it automatically finds the number of topics. The top vectors it creates are jointly embedded with the document and word vectors with distance between them representing semantic similarity.

## Results

The most frequent word in the TF-IDF analysis is “bitten,” followed by “flu,” “avian,” “fever,” “trap,” “fang,” “tail,” “hospital,” and “farm” (Figure a.). The main responses indicated that respondents were either bitten by a bat or rat when describing what type of accident they had regarding handling animals. “Avian” and “flu” refers to the participants response to name an animal transmitted disease. This was the only disease they mentioned in the survey. “Fever” refers to participants describing a specific symptom of diseases in general. “Trap” explains the participants’ specific job and “fang” and “tail” refer to the wildlife souvenirs they collected. “Hospital” is the answer respondents gave when discussing where they would go if they had symptoms of a disease related to handling animals. “Farm” describes one of the places where participants work.

**Figure a.**
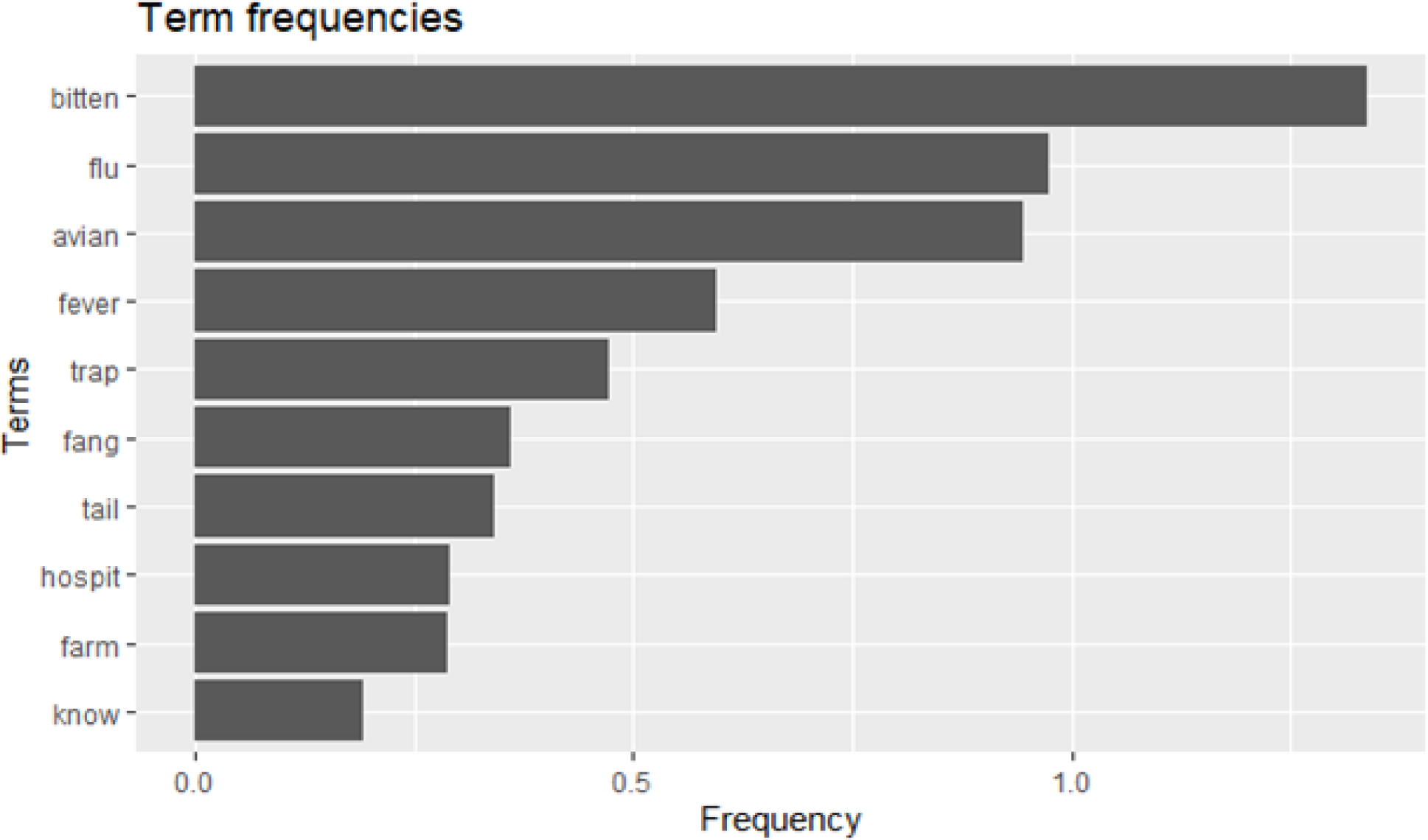
Term frequency results of the TF-IDF analysis TF-IDF analyzes individual word frequencies, we incorporated *ngrams* (Figure b.) which provides bigrams (two-word correlations) and trigrams (three-word correlations) to provide more context. Ngrams break down text into smaller components (tokens) consisting of *n* letters or words (Ganegedara, 2018).

**Figure b.**
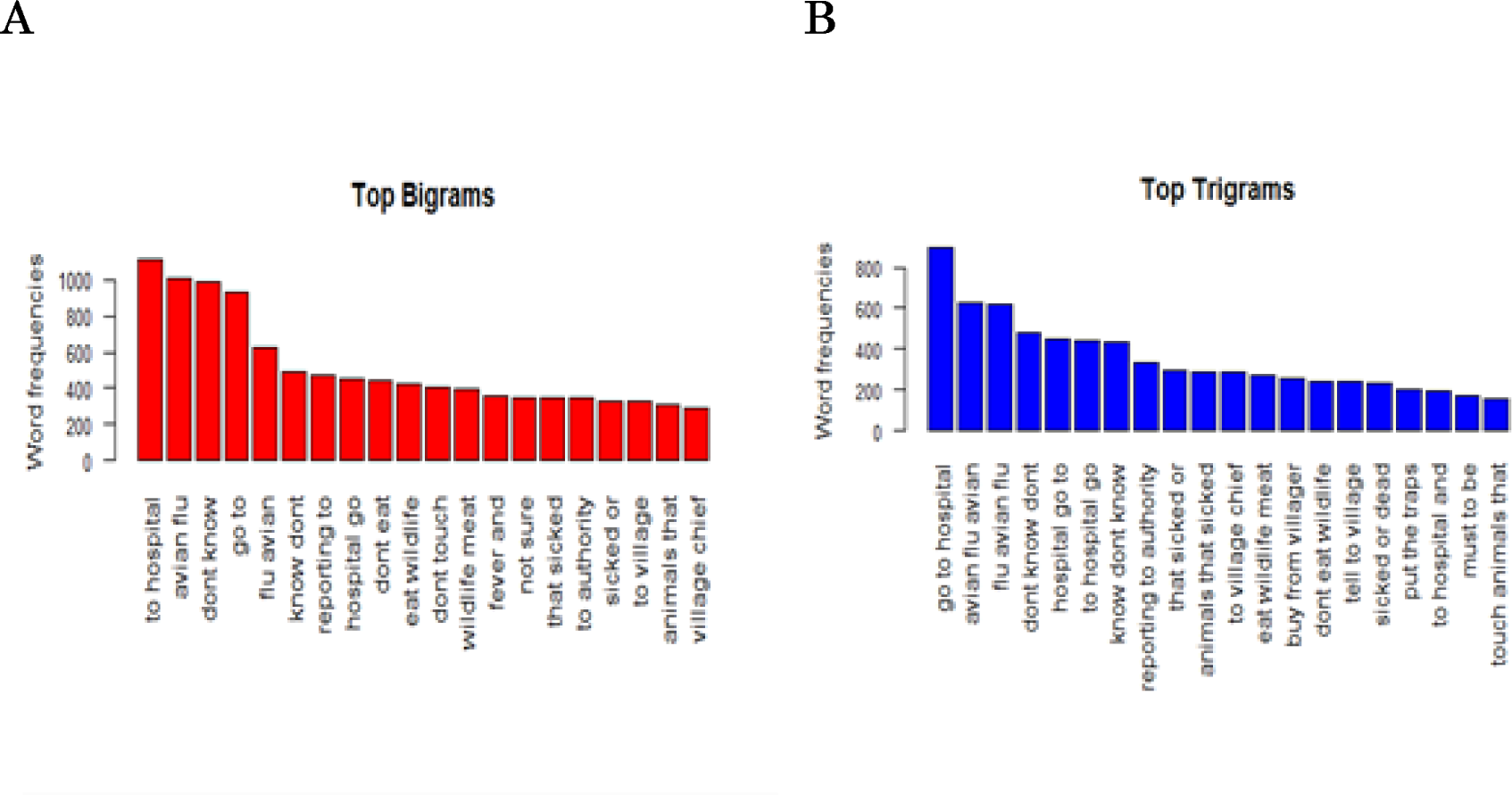
Results of the ngrams analyses A. Bigrams and B. Trigrams The LSA result s for the word “bat” indicated that “orchard” and “rodent” were related (Figure c.). In the LSA graphs below, words that are highly related to each other overlap in the graphs. This indicates that respondents could be encountering bats and rodents in their orchards. Bats are known to destroy fruit orchards around the world (Aziz et al., 2015). The word “rat” is related to “bats”, illustrating a relationship among potential food sources. Bat and rat could be related because participants interact with them frequently. “Eat” is related to “wildlife” and this it is related to the question asking them what they would do to prevent getting sick and they stated that “they don’t eat wildlife.” “Feces” is related to “trap” and “mist net” and it may mean that participants are finding and/or interacting with animal feces when they check their traps and mist nets.

**Figure c.**
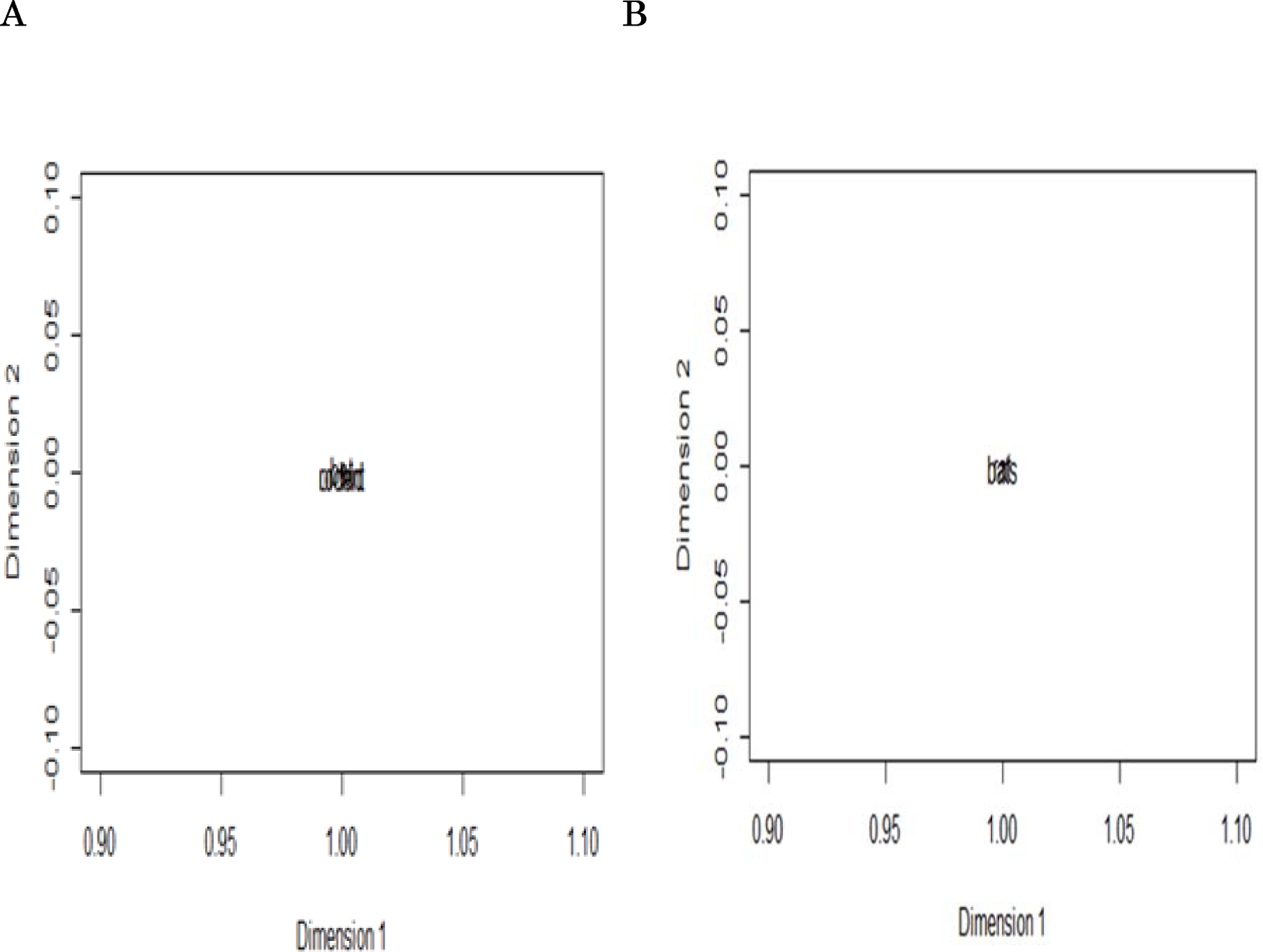
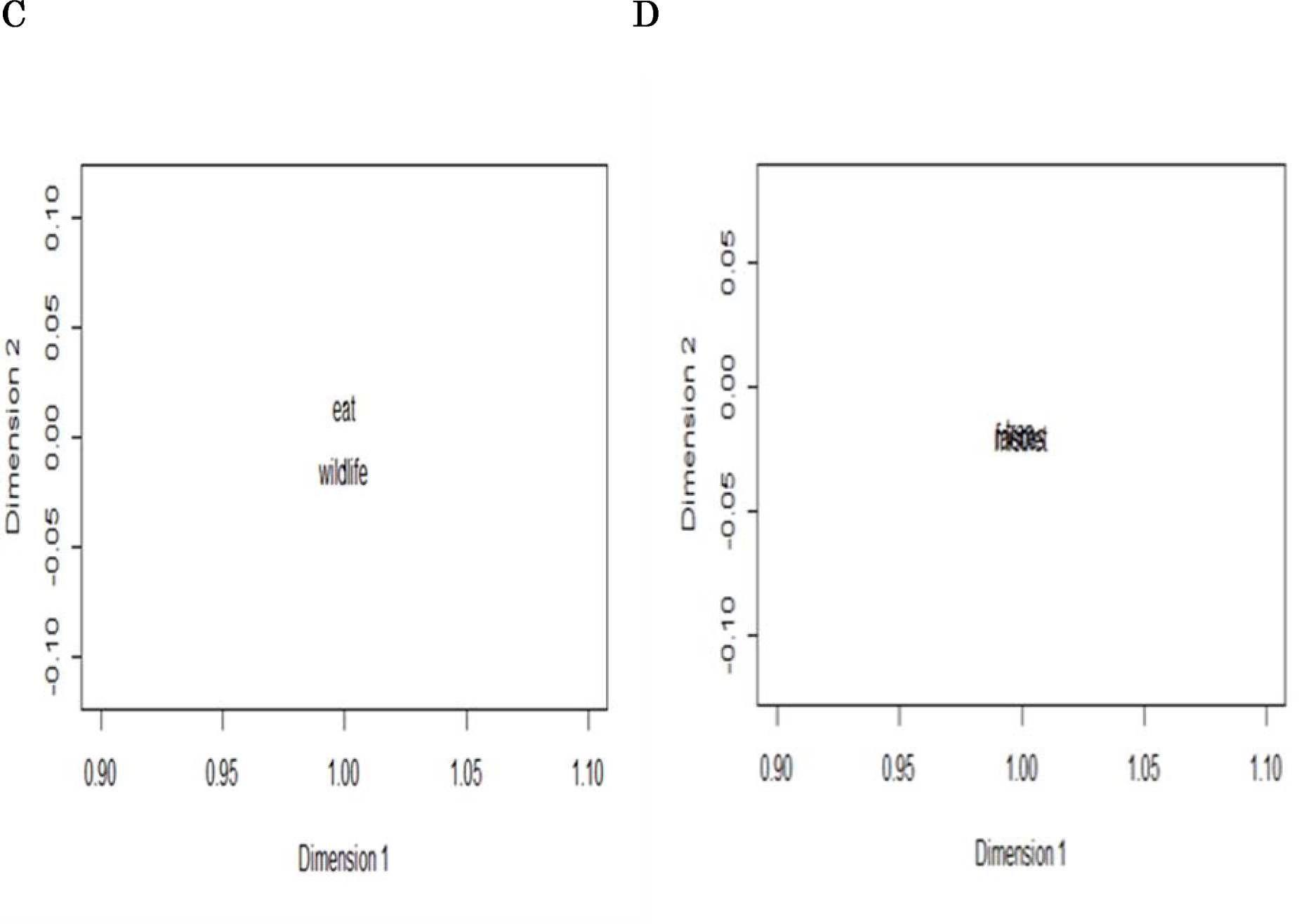
Latent Semantic Analysis *Note.* Results of the latent semantic analysis for terms that are similar including: A)“bat,” overlapping with “orchard” and “rat, B) “rat,” overlapping with “bat,” C) “eat,” overlapping with “wildlife,” and D) “feces” overlapping with the words “trap” and “mist net.”

The top bigrams include: “don’t know,” “avian flu,” “to hospital,” “to go,” “flu avian,” “don’t eat,” “know don’t,” and “eat wildlife” (Figure 1.3). The term “avian flu” is important and demonstrates that respondents understand it to be a zoonotic pathogen. Going “to hospital” is another frequent term that refers to respondents stating that that is what they would do if they had symptoms related to animal to human related diseases. The other terms are too vague to analyze. The top trigrams include: “go to hospital,” “avian flu,” “flu avian,” “don’t know don’t,”, “know don’t know,” and “to village chief” (Figure 1.3). “Reporting to authority” and “to village chief” refers specifically to whom respondents would tell if there were a zoonoses spillover event.

The 3 clusters in the k-means analysis include: Cluster 1-“avian flu,” and “bitten.” Cluster-2-“fever” and “hospital.” and Cluster 3-“rodent,” “pig,” “duck,” “hygiene,” and “medicine.” Cluster 1 relates to being protected from diseases by using/wearing protection as well as participants stating they w ere bitten by bat or rat. Cluster 2 relates to participants stating they would go to the hospital if they came down with a fever. Cluster 3 relates to being protected from animal to human diseases by being hygienic and seeking medicine when sick (Figure d.).

**Figure d.**
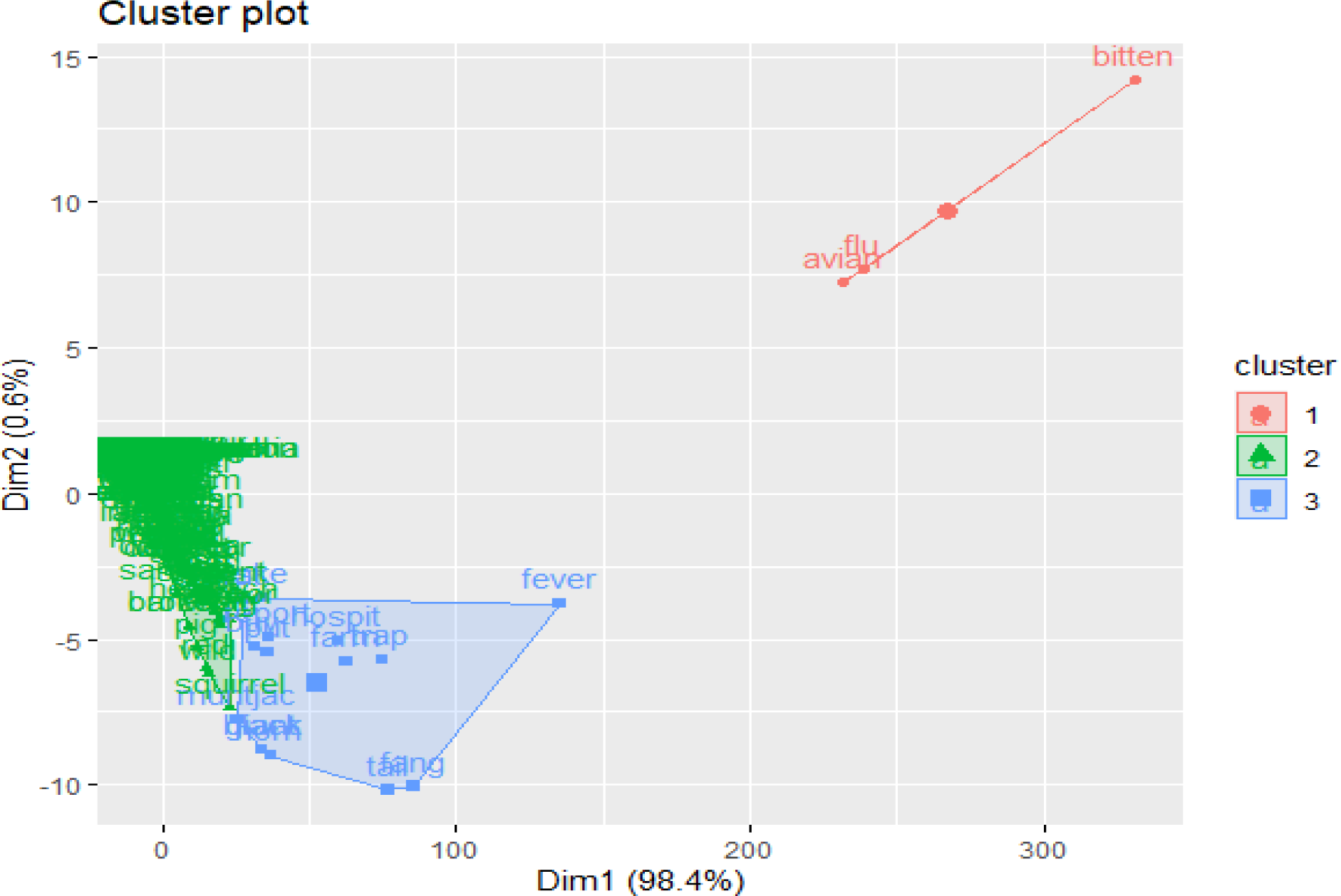
K-Means Cluster Plot *Note*. K-means cluster plot showing three different clusters: 1) “avian,” “flu” and “bitten” related to participants being aware of the only disease and also stating that they had been bitten by bat and rat, 2) relates to being protected from animal to human diseases by being “hygienic” and seeking “medicine” when sick, and relates to participants stating they would go to the “hospital” if they came down with a “fever”

Topic one for LDA is related to the survey question: what diseases are you aware of? (Figure e.). Participants stated they were aware of avian flu. Topic two is related to the survey question asking participants what their specific jobs were. Most stated they were farmers followed by trappers. Topic three is related to the question asking participants to identify their symptoms if they got sick. They answered that having a fever was one of the symptoms followed by being tired.

**Figure e.**
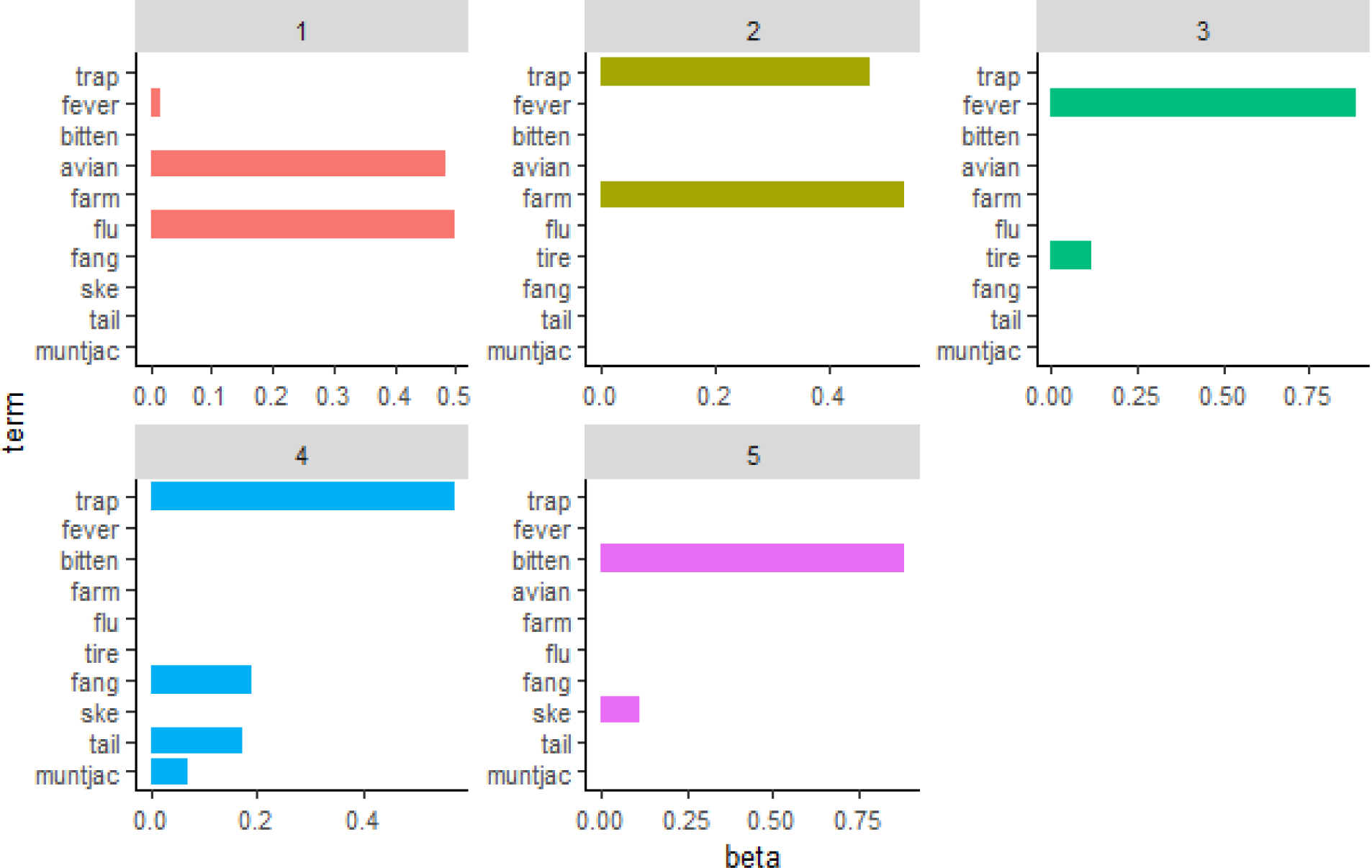
Latent Dirichlet Allocation *Note.* Top 5 topics for latent Dirichlet allocation model including: 1) “avian flu” showing participants’ knowledge of the only disease they are aware of, 2) “Trap” and “farm” describing where participants work, 3) “Fever” and “tired” describing participants’ awareness of disease symptoms, 4) “Trap,” “fang,” and “tail” showing that trappers keep wildlife fangs and tails as souvenirs, and 5) “Bitten” showing that participants were bitten by bat or rat.

Topic four is related to the question asking participants if they kept wildlife souvenirs. According to the LDA analysis, trappers kept wildlife fangs as souvenirs. Topic five shows that participants have been bitten by wildlife.

Top2Vec was used to find the top topics in the KAP survey. The analysis was done in Google Colab using Python 3.95. BERT (Devlin et al., 2018) sentence transformers were used to map sentences and paragraphs into a 768 dimensional dense vector space that can be used for semantic search (Rothman, 2021 & Ravichandaran, 2021).

The top topics are based on cosine similarities between words where 0 means there is now similarity and 1 represents the highest similarity. The top 5 topics for “bats” are “bat,” “banteng,” “bite,” “pest” and “chumpean.” (Figure f.). “Banteng” is a type of cattle in Cambodia and because it is related to the word “bat,” this could mean that banteng are susceptible to being bitten by bats. Some also see bats as a “pest.” “Chumpean” is a type of bird in Cambodia and this could be that they are in close proximity to “bats,” especially for those trappers who set mist nets. Some respondents did reply that they had been bitten by bats.

**Figure f.**
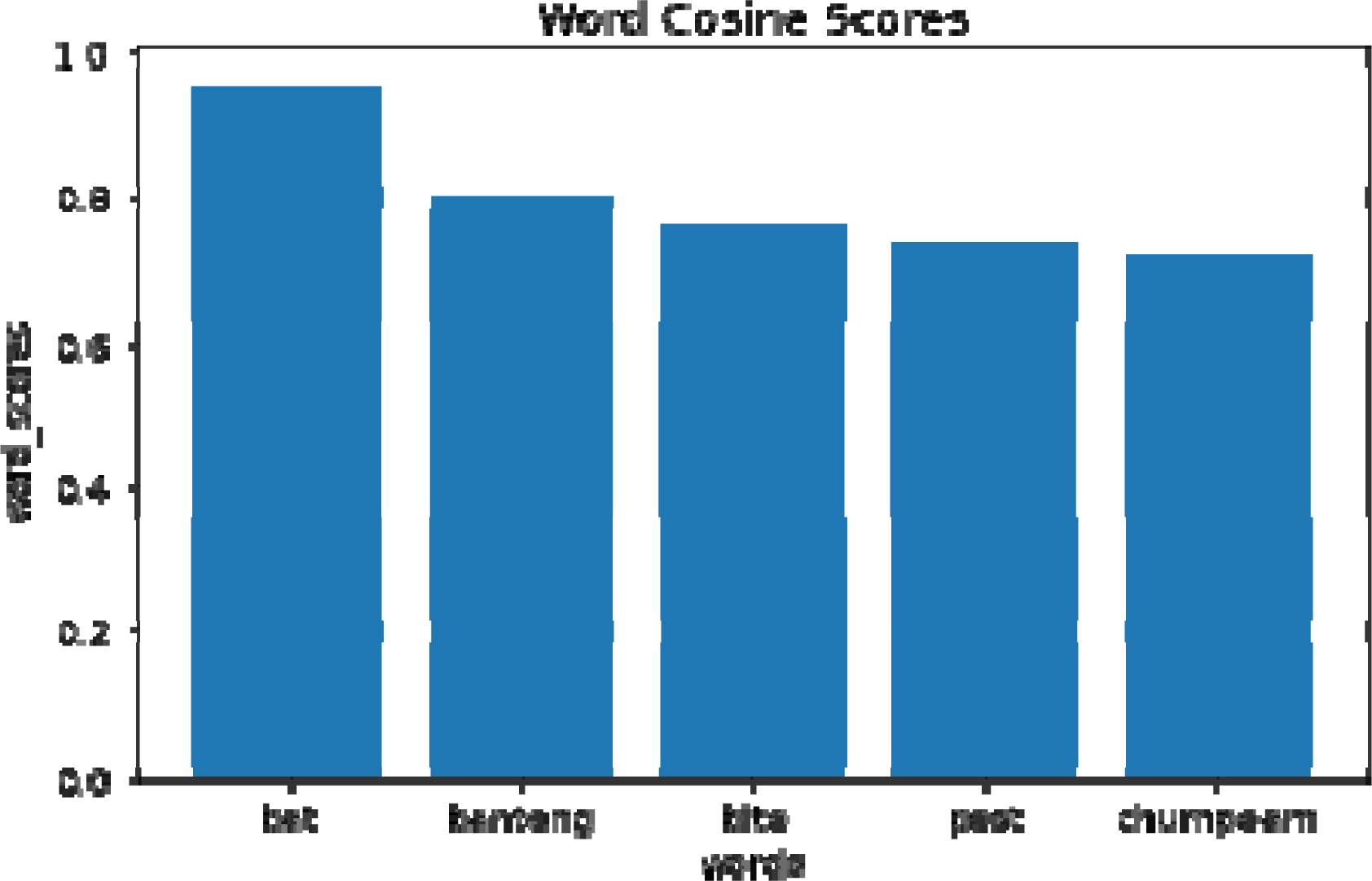
Top 5 Topics for “Bats,” are: “Bat,” “Banteng,” “Bite,” “Pest” and “Chumpeam.”

The top 5 topics for “rat” are: “rats,” “rodent,” “rabies,” “cage,” and “squirrel.” (Figure g.). Similarly, respondents stated that they know rats are pests, cause rabies, that they catch them and put them in cages, and that squirrels were like rats. Respondents also stated that they had been bitten by rat.

**Figure g.**
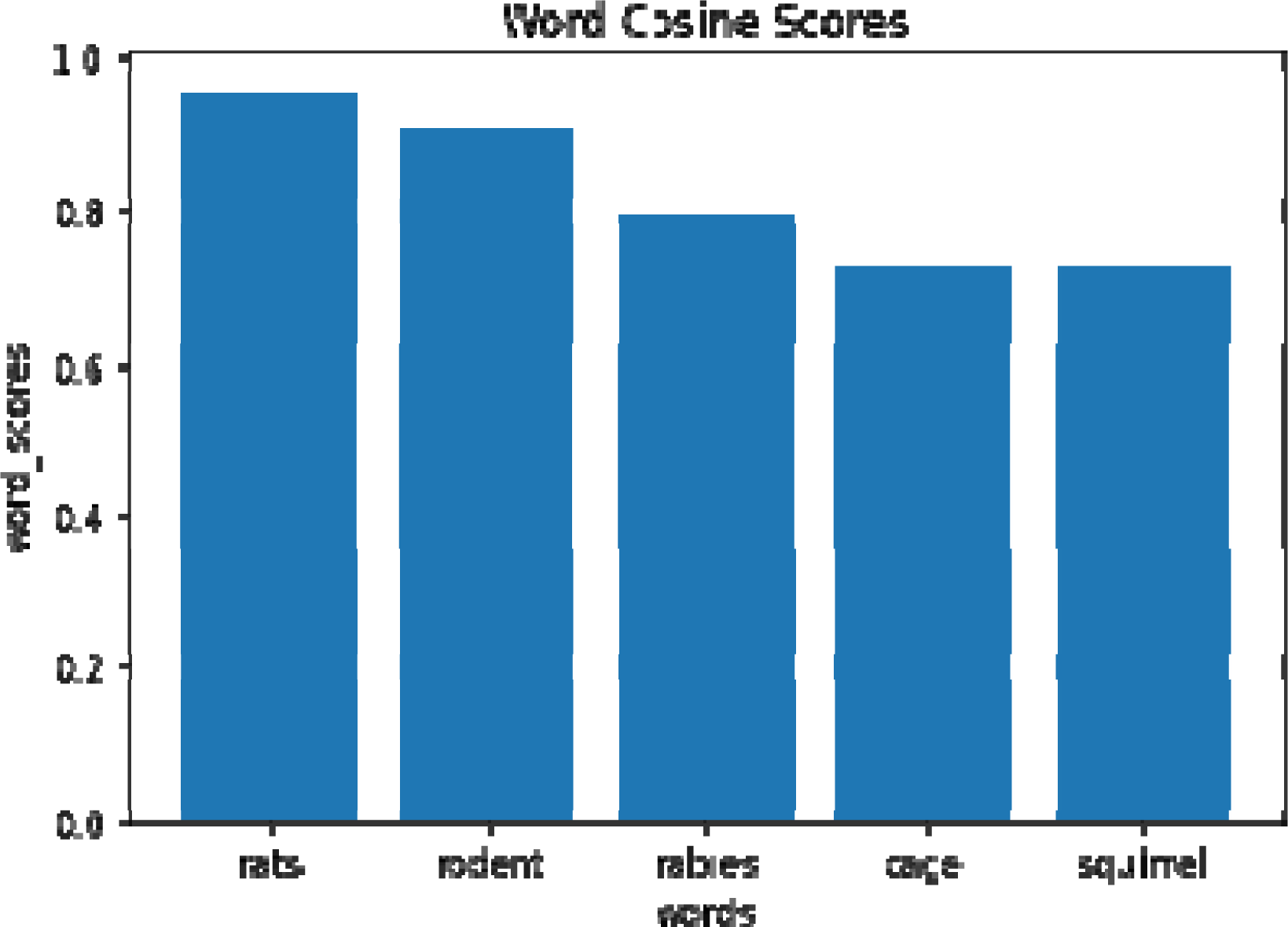
The Top 5 topics for “Rat” are: “Rats,” “Rodent,” “Rabies,” “Cage,” and “Squirrel.”

The top 5 topics for “hospital” are “patient,” “doctor,” “sanitary,” “medicine,” and “sanitized.” (Figure h.). Respondents here know to send their families and friends to the hospital if they get sick because they can help with medicine and injections.

**Figure h.**
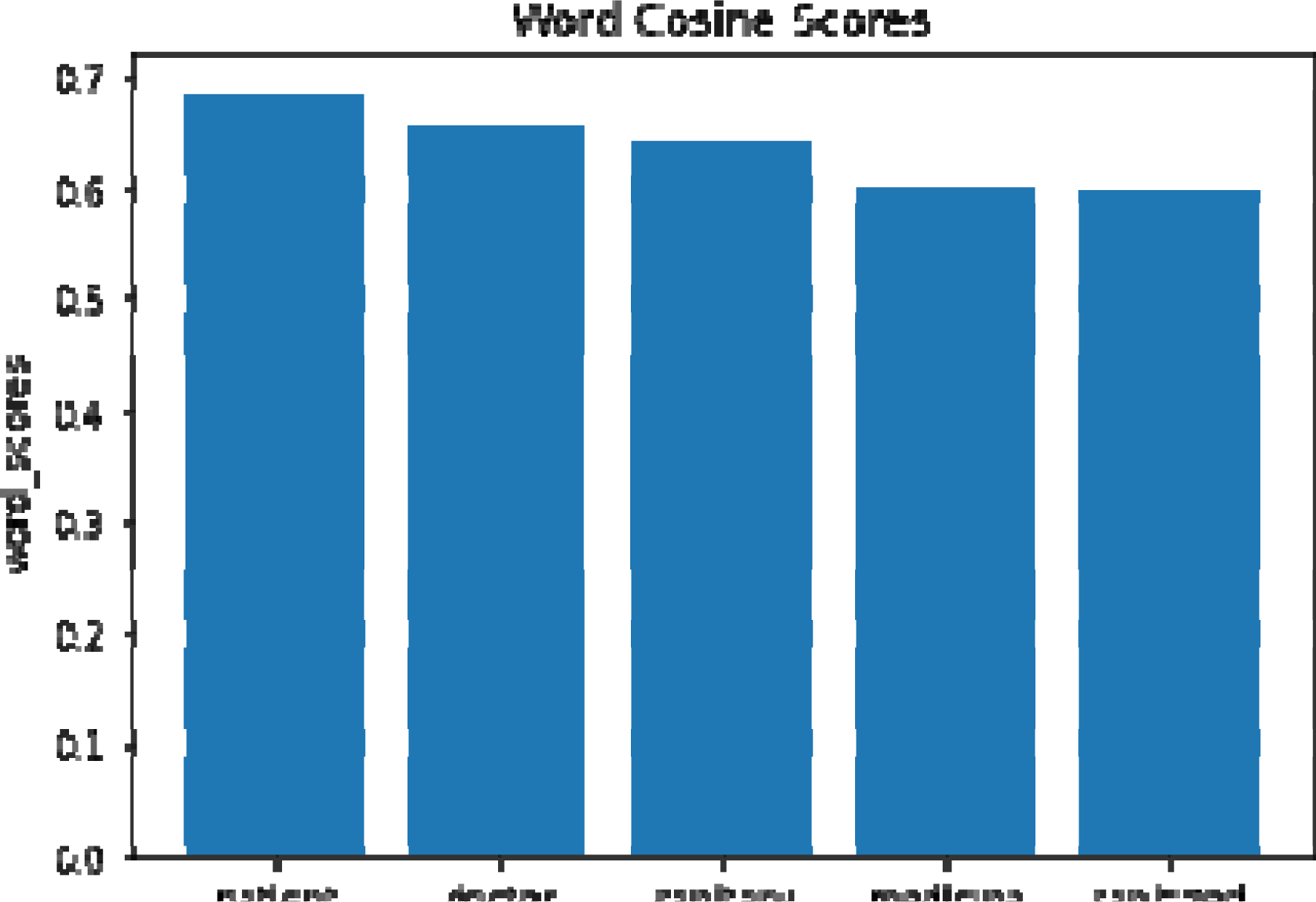
The Top 5 Topics for “Hospital” are: “Patient,” “Doctor,” “Sanitary,” “Medicine,” and “Sanitized.”

The top 5 topics for “flu” are “influenza,” “fever,” “dengue,” “ebola,” and “sick.” (Figure i.). Respondents are aware that flu is a symptom for possible diseases like dengue and Ebola and that it is a sign of being sick.

**Figure i.**
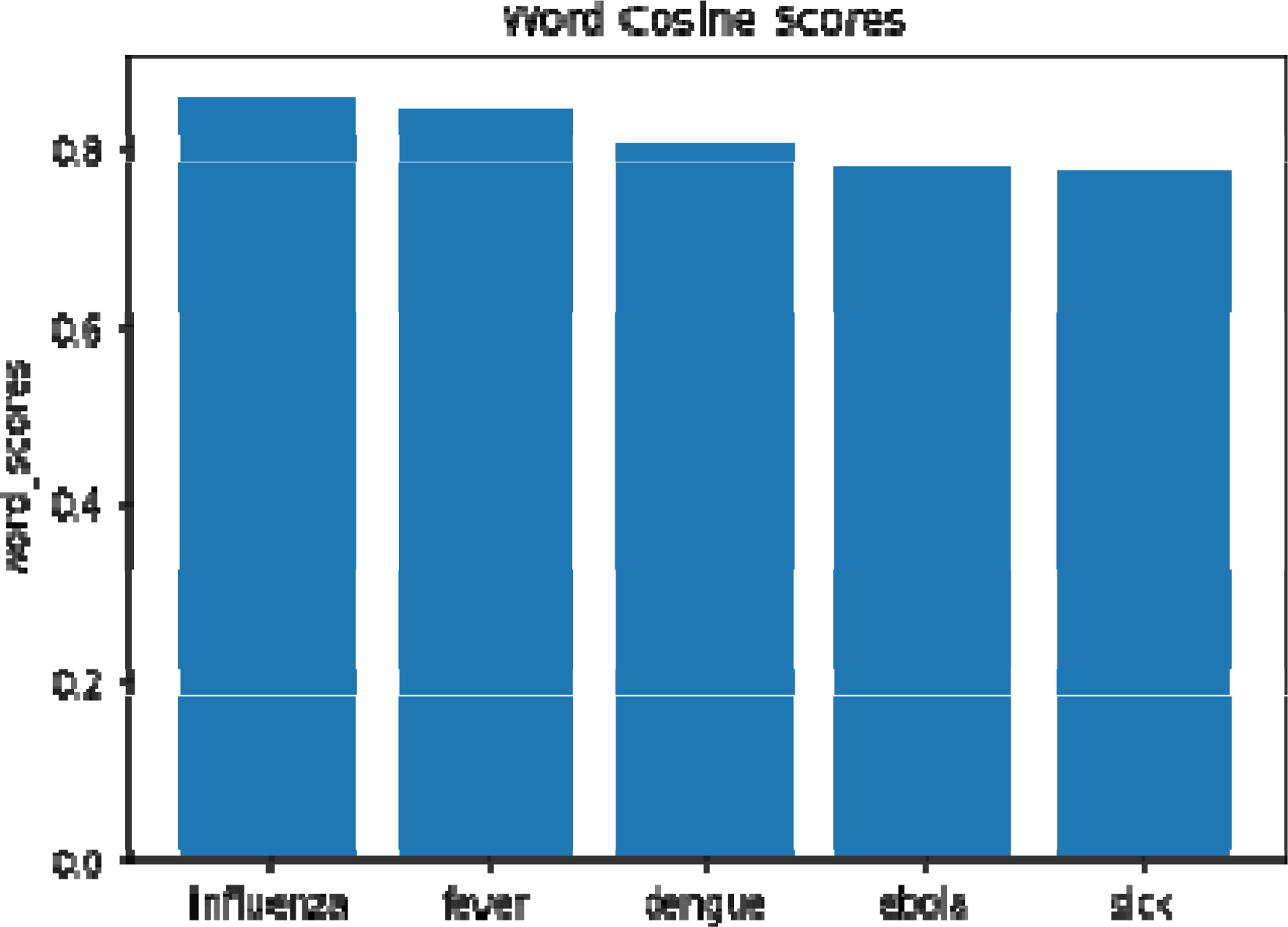
The Top 5 Topics for “Flu” are: “Influenza,” “Fever,” “Dengue,” “Ebola,” and “Sick.”

The top 5 topics for “sick” are “disease,” “flu,” “feverish,” “fever,” and “pest.” (Figure j.). Respondents here are aware of the symptoms of being sick related to wildlife diseases. Many respondents stated that they got bitten by “pests” such as bat and/or rat.

**Figure j.**
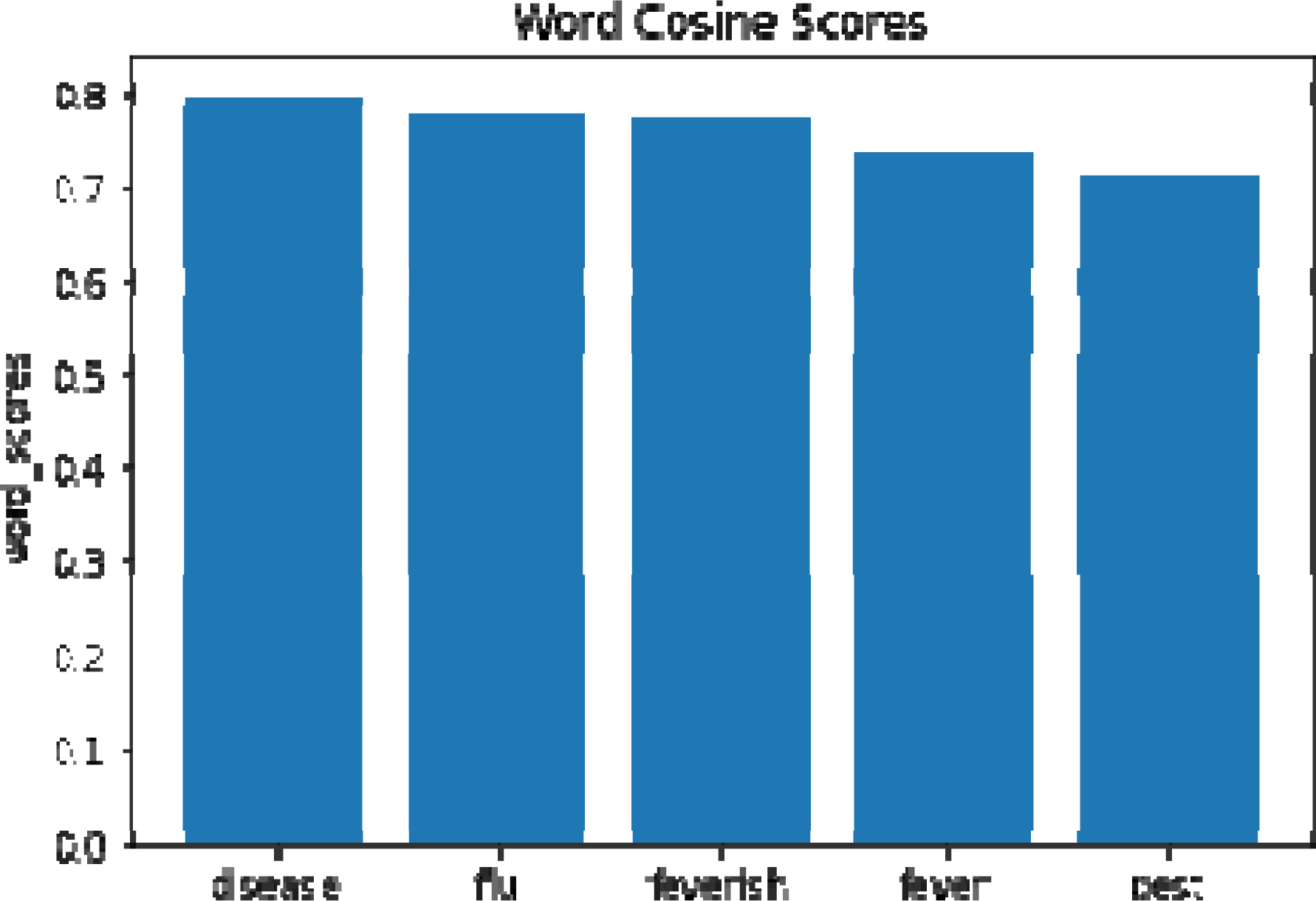
The Top 5 Topics for “Sick” are: “Disease,” “Flu,” “Feverish,” “Fever,” and “Pest.”

## 1.5 Discussion

Monagin et al. (2018) found that handling wildlife increases the likelihood of showing seropositivity for several pathogens like Hantavirus, SARS CoV, or SFTS bunyavirus. Participants in our study stated they handled or encountered wildlife directly since they set wildlife traps and mist nets. The NLP analyses also show that respondents were bitten by wildlife and most stated that they were either bitten by a bat or rat. Wang & Anderson (2019) state that because of the diversity of bats including their geographic diversity, that they host and serve as a reservoir for many viruses that they can directly transmit to humans either by being bitten, butchering them, or working with their manure. Some of these diseases include: severe respiratory syndrome coronavirus (SARS-CoV), Hendra virus (HeV), Menangle virus (MenPV), and Nipah virus (NiV).

Luis et al. (2014) found in their study that rats were just as serious as bats as zoonotic reservoirs that transmitted infectious diseases to humans. Monagin et al. (2018) found that increased contact with rats increased the likelihood of seropositivity which are popular in open-air markets. NLP modeling above also showed that participants were aware of avian flu.

In their study investigating household practices and zoonoses transmission in Cambodia, Osbjer et al. (2015) found that over 65% of respondents were aware that avian flu is a transmissible disease. The researchers state that this was the only zoonotic disease they were aware of because of a countrywide avian flu awareness campaign that Cambodia had initiated previously.

Another NLP finding showed that participants were aware that if they had a fever or headache that they should go to the hospital and that they should use hygiene and medicine in case they get sick. However, being bitten was not mentioned when describing when they should go to the hospital. Nevertheless, Suttie et al. (2018) in their research of avian flu in chickens and ducks in Cambodia found that reporting of avian flu symptoms was minimal and that compensation to Cambodian owners of chickens and ducks were non-existent. Thus, respondents could have stated to interviewers that they should go to the hospital and use hygiene and medicine to look favorable in front of interviewers. In Monagin et al. (2018), respondents could have underreported their practices regarding handling wildlife since it is illegal in China. Osbjer et al. (2015) similarly state that respondents might over-report good hygiene practices due to intentionally or unintentionally perceived desirable responses.

Because of the participants’ details discovered in the NLP analyses above, recommendations include: 1) health officials need to educate participants to wear protective gear to prevent from being bitten by bats and rats during their jobs with these animals, and 2) health officials need to educate participants about the danger of different types of zoonotic diseases including Ebolavirus, Mojianvirus, etc., so that these participants can recognize the risks when handling bats and rats, and so they can take early action by seeking medical help as soon as they are bitten. Osbjer et al. (2015) study showed that the avian flu awareness campaign in Cambodia was successful as far as knowledge about it are concerned while awareness of other zoonotic diseases were unknown. Thus, it is possible to have a zoonotic awareness campaign countrywide in Cambodia detailing how to protect themselves from bats and rats and which diseases they transmit to humans which can be harmful so that Cambodians take necessary precautions.

## Data Availability

All data produced in the present work are contained in the manuscript.

## Declarations

### Ethical Approval

Consent was provided by all participants to participate in the study and allow the work to be published as long as data was anonymized. This study was approved and undertaken by the Forestry Administration in Cambodia from 2014 to 2017. Their International Research Development Centre grant number is 106915-001.

### Author Contributions

Roger Geertz Gonzalez did 100% of the data cleaning, analysis, write up, creating tables and figures, and writing the conclusion.

### Availability of data and materials

Data is available upon reasonable request.

